# Acute hemodynamic effects after Impella 5.5 in cardiogenic shock and association with clinical outcomes

**DOI:** 10.64898/2026.05.19.26353572

**Authors:** Gabrielle Daso, Paridhi Gupta, Joseph J Locascio, Van-Khue Ton, Erin Coglianese, Kamila Drezek, Joyce W Wald, Eriberto Michel, David A D’Alessandro, Bin Q Yang

**Affiliations:** Department of Internal Medicine, Massachusetts General Hospital, Boston MA; Division of Cardiology, Massachusetts General Hospital, Harvard Medical School, Boston MA; Department of Neurology, Harvard Medical School, Boston MA; Division of Cardiac Surgery, Massachusetts General Hospital, Harvard Medical School, Boston MA; Division of Cardiology, University of Pennsylvania, Philadelphia PA

**Keywords:** Impella 5.5, cardiogenic shock, right ventricular function, pulmonary artery hemodynamics

## Abstract

Cardiogenic shock (CS) is associated with high short-term mortality and the use of temporary mechanical circulatory support (tMCS) devices, especially left-sided microaxial flow pumps (Impella, Abiomed), has increased in recent years. However, few studies have investigated tMCS’s effect on right ventricular-pulmonary artery (RV-PA) hemodynamics and its impact on clinical outcomes. We retrospectively analyzed all adult patients implanted with Impella 5.5 at our institution with acute myocardial infarction or acute decompensated heart failure-induced CS between 2019 to 2023. We found that Impella 5.5 led to an early improvement in RV-PA hemodynamics, even in patients with poor baseline RV function. In addition, we found that RV function itself did not predict death, post-heart transplant right ventricular-primary graft dysfunction, or post-left ventricular assist device severe RV failure. However, an increase in right atrial:pulmonary capillary wedge pressure ratio (RA/PCWP) despite tMCS support was a powerful prognosticator. Our study sheds important insight into anticipated hemodynamic changes after Impella 5.5 placement, supports the use of early tMCS even in patients with marginal RV function in the setting of left heart disease, and highlights the importance of serial assessment of RA/PCWP as a key determinant of CS outcomes.

## Introduction

Cardiogenic shock (CS) is a clinical syndrome characterized by hypotension and end-organ hypoperfusion, associated with short-term mortality rate ranging from 30% to >50%.^1–4^ Early studies have focused on CS due to acute myocardial infarction (AMI-CS), although the prevalence of acute decompensated heart failure resulting in CS (HF-CS) is rising.^1,5,6^ Right ventricular (RV) function is increasingly recognized as a key determinant of CS outcomes, as RV dysfunction is present in roughly 30% of patients with CS and is an independent predictor of death and hospitalization.^7,8^ Few studies to date have examined detailed changes in RV hemodynamics in patients with CS, especially after the initiation of left-sided temporary mechanical circulatory support devices (tMCS).

Early hemodynamic support in CS can prevent downstream consequences such as end-organ dysfunction, inflammation, and systemic vasodilation (cardiometabolic shock).^1,9–11^ As such, the use of percutaneous microaxial flow pumps (Impella, Abiomed) has increased in frequency. The DanGer Shock trial showed a mortality benefit in patients treated with Impella CP in AMI-CS; however, patients with ADHF-CS and severe pre-existing RV dysfunction were excluded.^12^ Impella 5.5 is a surgically-implanted pump that can provide up to 5.5L/min of cardiac output and has been increasingly utilized to bridge CS patients to recovery or heart replacement therapies such as durable left ventricular assist devices (LVAD) or heart transplant (HT), especially after the 2018 HT allocation system change.^5,6^

Despite providing robust left ventricular (LV) support, one clinical concern about the use of Impella 5.5 is RV response to the increased preload. Yet, Impella 5.5 may also help RV function by reducing pulmonary artery (PA) and pulmonary capillary wedge pressures (PCWP) and RV afterload. Given the equipoise, a detailed understanding of how Impella 5.5 affects RV and PA hemodynamics across a range of baseline RV functions can provide valuable information to help select appropriate candidates for tMCS. As such, our study was designed to investigate acute hemodynamic changes after Impella 5.5 placement in patients with AMI-CS and HF-CS. We also sought to investigate which hemodynamic parameters are associated with clinical outcomes. We hypothesized that even in patients with poor RV function, Impella 5.5 placement leads to a net benefit by preferentially reducing RV afterload, enhancing RV-PA coupling, and improving end-organ perfusion.

## Materials & Methods

### Study Design and Population

This was a retrospective single-center analysis of all patients over the age of 18 at Massachusetts General Hospital who received Impella 5.5 for AMI-CS and HF-CS between 2019 to 2023. Patients were excluded if they received Impella 5.5 before high-risk procedures (coronary intervention or ventricular tachycardia ablation), post-cardiotomy shock, underwent concurrent extracorporeal membrane oxygenation (ECMO) cannulation or RV tMCS, or did not have pulmonary artery catheter for hemodynamic monitoring. Patients were further stratified based on their baseline RV function, determined by a combination of tricuspid annular plane systolic excursion (TAPSE) and right atrial:PCWP ratio (RA/PCWP).^13^ “Good” RV function was defined as TAPSE ≥ 16 mm and RA/PCWP ≤ 0.6 whereas “bad” RV function was characterized by TAPSE < 16mm and RA/PCWP > 0.6. Patients who met 1 of the 2 criteria were classified as “intermediate.”

### Data collection

Demographic and clinical data were collected from electronic medical records at baseline (pre-implantation) and at 12, 24, and 48 hours after Impella 5.5 placement. Specifically, we collected serial hemodynamic measurements, laboratory markers of end-organ perfusion, and clinical status such vasopressor-inotrope score (VIS) and Society for Cardiovascular Angiography & Interventions (SCAI) staging.^3,4^ As some patients did not have PCWP measured at the exact aforementioned time points, we extrapolated the closest documented PCWP and in cases of missing data, assumed a constant PA diastolic pressure gradient to estimate the PCWP.^14^

### Outcomes

The main objective was to analyze trends of right atrial pressure (RAP), RA/PCWP, pulmonary artery pulsatility index (PAPi), RV stroke work index (RVSWI), and pulmonary arterial elastance (Ea) as markers of RV and PA hemodynamics before and after Impella 5.5 implantation.^13,15^ The primary clinical outcome was death during index hospitalization, censoring those who received durable LVAD or HT as favorable events. The secondary outcome was a composite of death during index hospitalization, post-HT RV-primary graft dysfunction (PGD), or INTERMACS-defined severe RV failure post-LVAD.

### Statistical analysis

Demographic data were analyzed with descriptive statistics. Continuous variables were characterized as mean or median with standard deviation and interquartile range, as appropriate. Categorical variables were detailed using percentage of total cohort. SAS statistical software (version 9.4) was used for the analyses.

We fit mixed effects longitudinal models to analyze the change over time in hemodynamic parameters, markers of hypoperfusion, and hepatorenal function, respectively. Each model was adjusted for age and sex as fixed effects and stratified by RV function to assess group-specific trajectories. The model for hepatorenal function was also stratified by the presence of underlying chronic kidney disease (CKD). Time was measured as linear and quadratic variables to account for dynamic and differential changes in rate, resulting in curvilinear plots. Higher order fixed and random terms were pretested and removed from the model if nonsignificant to increase power for effects of more interest.

We subsequently generated a multivariable logistic regression model adjusted for age, baseline RV function, and the rate of change in hemodynamic variables to predict the primary and secondary clinical outcomes. Due to the sample size, we limited the primary model to these main variables to avoid overfitting. We then performed pre-specified sensitivity analyses, further adjusting for sex and linear regression of the hemodynamic variables to better capture the overall rate of change across the entire 48-hour window post-Impella 5.5.

## Results

### Baseline clinical characteristics

Between 12/2019 and 12/2023, 179 patients underwent Impella 5.5 implantation. After exclusions, our final cohort consisted of 86 patients, with 21% (N=18) categorized as having good, 56% (N=48) intermediate, and 23% (N=20) bad RV function (**Supplemental Figure 1**). Baseline characteristics are presented in **Table I**. Patients with intermediate RV function were younger (p=0.043) but there were no significant differences in sex, race, or ethnicity compared to the other groups. Patients with bad RV function had significantly higher rates of atrial fibrillation, chronic kidney disease, and lower sodium and estimated glomerular filtration rate (eGFR).

**Table 1:** Baseline characteristics of patients who underwent Impella 5.5 implantation, stratified by baseline right ventricular (RV) function. eGFR=estimated glomerular filtration rate (by creatinine clearance), INR=international normalized ratio, MELD=model for end-stage liver disease, IABP=intra-aortic balloon pump, LVAD=left ventricular assist device, PGD=primary graft dysfunction.

| Baseline Characteristics | Good<br>(18, 21%) | Intermediate<br>(48, 56%) | Bad<br>(20, 23%) | P-value<sup>2</sup> |
| --- | --- | --- | --- | --- |
| Age (years)<sup>1</sup> | 64 ± 11 | 56 ± 12 | 62 ± 9.1 | 0.043 |
| Male sex, %yes (n) | 13 (72) | 35 (73) | 16 (80) | 0.843 |
| Body mass index (kg/m<sup>2</sup>)<sup>1</sup> | 27 ± 5.2 | 30 ± 7.1 | 28 ± 6.0 | 0.186 |
| Impella duration in days<br>(median ± SD) | 9 ± 9.5 | 12 ± 15 | 13 ± 29 | 0.218 |
| **Race** |  |  |  | 0.089 |
| White | 15 (83) | 32 (67) | 14 (70) |  |
| Black | 0 (0) | 7 (15) | 3 (15) |  |
| Asian | 3 (17) | 2 (4) | 0 (0) |  |
| Multiracial/unknown | 0 (0) | 7 (15) | 3 (15) |  |
| Hispanic ethnicity, %yes<br>(n) | 3 (17) | 2 (4) | 3 (15) | 0.317 |
| **Comorbidities, %yes (n)** |  |  |  |  |
| Ischemic cardiomyopathy | 8 (44) | 20 (42) | 10 (50) | 0.803 |
| Hypertension | 9 (50) | 26 (54) | 11 (55) | 0.958 |
| Diabetes mellitus | 6 (33) | 16 (33) | 8 (40) | 0.909 |
| Atrial fibrillation | 3 (17) | 17 (35) | 15 (75) | <0.001 |
| Ventricular tachycardia | 5 (28) | 8 (17) | 5 (25) | 0.486 |
| Stroke | 0 (0) | 5 (10) | 1 (5) | 0.534 |
| Chronic kidney disease | 5 (28) | 12 (25) | 12 (60) | 0.022 |
| **Laboratory Values<sup>1</sup>** |  |  |  |  |
| Sodium (mmol/L) | 136±4.7 | 134±4.6 | 132±5.6 | 0.020 |
| Blood Urea Nitrogen<br>(mg/dL) | 36±18 | 32±17 | 46±17 | 0.010 |
| Creatinine (mg/dL) | 1.66±0.94 | 1.73±0.91 | 2.2±1.0 | 0.158 |
| eGFR (mL/min/1.73m<sup>2</sup>) | 58±31 | 55±28 | 38±18 | 0.012 |
| Total bilirubin (mg/dL) | 1.7±1.4 | 1.4±1.1 | 1.8±1.4 | 0.387 |
| Albumin (g/dL) | 3.3±0.45 | 3.5±0.51 | 3.5±0.39 | 0.261 |
| INR | 1.4±0.36 | 1.4±0.35 | 1.7±0.94 | 0.515 |
| NT-proBNP (pg/dL) | 9624±8235 | 9050±10208 | 23749±57356 | 0.605 |
| Lactate (mmol/L) | 2.3±2.8 | 2.7±2.5 | 2.6±3.9 | 0.911 |
| MELD Score | 15±8.2 | 15±7.1 | 20±9.0 | 0.114 |
| **Prior mechanical circulatory support device** |  |  |  | 0.081 |
| IABP | 4 (22) | 3 (6) | 2 (10) | 0.174 |
| Impella CP | 2 (11) | 3 (6) | 3 (15) | 0.466 |
| **Outcomes** |  |  |  |  |
| Heart transplant | 5 (28) | 20 (42) | 10 (50) | 0.400 |
| LVAD | 5 (28) | 8 (17) | 3 (15) | 0.529 |
| Composite outcome | 4 (22) | 22 (46) | 5 (25) | 0.109 |
| Death | 2 (11) | 16 (33) | 4 (20) | 0.154 |
| Post-transplant PGD-RV | 0 (0) | 1 (2) | 0 (0) | 1.000 |
| Post-LVAD RV failure | 2 (11) | 5 (10) | 0 (0) | 0.674 |

<sup>1</sup>Mean ± SD; (n, %)
<sup>2</sup>One-way analysis of means (not assuming equal variances); Fisher's exact test

At time of Impella 5.5 placement, 91% of patients presented in SCAI stage C or greater CS despite high pharmacological support. There were no significant differences in mean arterial pressure, LV ejection fraction, LV end-diastolic diameter, cardiac output and index, and VIS between the groups. Hemodynamically, patients with bad RV function had higher RAP (p<0.001), higher RA/PCWP (p=0.009), and lower PAPi (p<0.001) despite similar mean PA pressures (p=0.605), pulmonary vascular resistance (p=0.962), and Ea (p=0.277). Patients with bad RV function also had significantly lower heart rate (85 vs. 98 vs. 100, p=0.015, **Table II**).

**Table 2:** Society for Cardiovascular Angiography & Interventions (SCAI) Staging, echocardiographic, and hemodynamic data at time of Impella 5.5 placement, stratified by baseline right ventricular (RV) function. LV=left ventricular, TAPSE=tricuspid annular planar systolic excursion (mm), RAP=right atrial pressure, PA=pulmonary arterial, PCWP=pulmonary capillary wedge pressure, CO=cardiac output, CI=cardiac index, PVR=pulmonary vascular resistance.

|  | **Good**<br>**(18, 21%)** | **Intermediate**<br>**(48, 56%)** | **Bad**<br>**(20, 23%)** | **P-value<sup>2</sup>** |
| --- | --- | --- | --- | --- |
| **SCAI Stage, n (%)** |  |  |  | 0.825 |
| A | 0 (0) | 2 (4) | 2 (10) |  |
| B | 0 (0) | 4 (8) | 0 (0) |  |
| C | 11 (61) | 23 (48) | 11 (55) |  |
| D | 5 (28) | 12 (25) | 4 (2) |  |
| E | 2 (11) | 7 (15) | 3 (15) |  |
| **Hemodynamics<sup>1</sup>** |  |  |  |  |
| Systolic Blood Pressure (mmHg) | 104±15 | 102±18 | 104±12 | 0.855 |
| Diastolic Blood Pressure (mmHg) | 59±12 | 60±9.8 | 60±9.7 | 0.531 |
| Mean Arterial Pressure (mmHg) | 75±7.1 | 76±14 | 77±8.3 | 0.881 |
| Heart Rate (bpm) | 100±20 | 98±26 | 85±16 | 0.015 |
| Vasoactive-Inotropic Score | 976±1774 | 1040±2019 | 869±1550 | 0.933 |
| **Transthoracic Echocardiogram<sup>1</sup>** |  |  |  |  |
| LV Ejection Fraction (%) | 22±6.6 | 21±8.6 | 18±5.9 | 0.198 |
| LV End-Diastolic Diameter (mm) | 65±9.8 | 62±9.6 | 61±19 | 0.548 |
| TAPSE (mm) | 20±1.8 | 15±3.8 | 13±2.9 | <0.001 |
| RV Systolic Pressure (mmHg) | 47±15 | 46±9.5 | 41±8.9 | 0.168 |
| **Right Heart Catheterization<sup>1</sup>** |  |  |  |  |
| RAP (mmHg) | 10±4.4 | 13±6.8 | 18±4.7 | <0.001 |
| PA Systolic Pressure (mmHg) | 54±14 | 50±15 | 51±12 | 0.585 |
| PA Diastolic Pressure (mmHg) | 26±7.6 | 25±10 | 26±7.3 | 0.995 |
| Mean PA Pressure (mmHg) | 37±7.1 | 35±11 | 35±9.0 | 0.605 |
| PCWP (mmHg) | 31±7.0 | 25±9.5 | 23±7.1 | 0.009 |
| CO by Thermodilution (L/min) | 3.7±1.6 | 3.9±1.1 | 4.2±1.7 | 0.715 |
| CI by Thermodilution (L/min/m<sup>2</sup>) | 2.1±0.81 | 1.9±0.45 | 2.1±0.84 | 0.578 |
| CO by Fick (L/min) | 4.9±2.4 | 4.2±1.3 | 3.9±1.3 | 0.397 |
| CI by Fick (L/min/m<sup>2</sup>) | 2.7±1.4 | 2.0±0.51 | 1.9±0.56 | 0.265 |
| PVR (Wood units) | 3.7±5.7 | 3.4±2.4 | 3.5±2.3 | 0.962 |
| RA/PCWP Ratio | 0.36±0.14 | 0.54±0.26 | 0.81±0.17 | <0.001 |
| PA pulsatility Index | 28±14 | 25±9.3 | 25±8.3 | <0.001 |
| RV Stroke Work Index (g·m/m<sup>2</sup>) | 7.5±4.6 | 6.3±4.9 | 5.6±2.5 | 0.346 |
| PA Elastance (mmHg/mL) | 1640±853 | 1263±601 | 1273±630 | 0.277 |

<sup>1</sup>Mean ± SD; (n, %)
<sup>2</sup>One-way analysis of means (not assuming equal variances)

### Acute hemodynamic change after Impella 5.5

After Impella 5.5 implantation, we found significant changes in hemodynamics over time, peaking between 30-36 hours post-implantation. RAP decreased in all patients, although the greatest effect was seen in patients with bad RV function (p<0.001, **Figure 1a**). We noted a paradoxical increase in RA/PCWP across all RV groups that differed by sex with females having a much greater rise compared to males (**Supplemental Figure 2**). Measures of intrinsic RV function such as PAPi and RVSWI improved in all patients but those with good baseline RV function experienced the greatest benefit, resulting in doubling of PAPi and 50% increase in RVSWI, respectively (p<0.05, **Figure 1b-c**). Ea decreased in all cohorts, although patients with good RV function had a greater magnitude of change as compared to those with bad or intermediate RV function (p=0.05, **Figure 1d, Supplemental Figure 3**).

**Figure 1:**
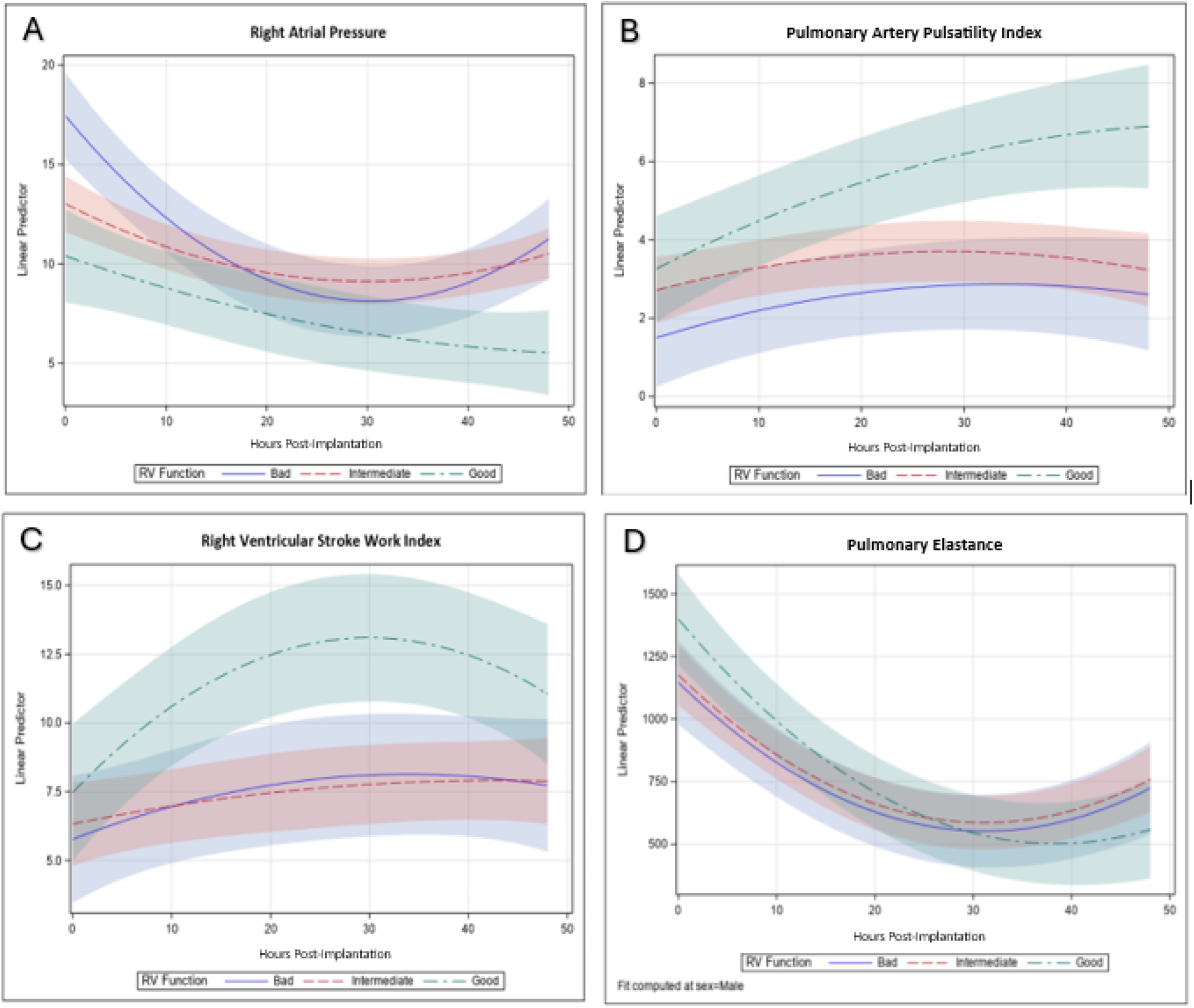
Mixed effects longitudinal models of changes in right ventricular (RV) and pulmonary artery hemodynamics after Impella 5.5 placement, stratified by good, intermediate, and bad RV function. Panel D is specific to males; the corresponding curves for females are slightly steeper and is depicted in Supplementary Figure 2. Bands indicate 95% confidence intervals.

### Markers of end-organ function

In addition to the overall improvement in hemodynamics, end-organ perfusion markers improved after Impella 5.5. Renal function improved in all patients. Those with pre-existing CKD demonstrated a greater rise in eGFR at a mean linear rate of 0.88 units/hour as compared to those with normal baseline kidney function (**Figure 2a**, p<0.05). Lactate and VIS both decreased, consistent with restoration of cardiac output and improved tissue perfusion. Interestingly, older patients had worse renal function and higher lactate at baseline and their response to tMCS was attenuated (p<0.01, respectively, **Figure 2b-c**). We did not observe a meaningful change in MELD between baseline and 48 hours after Impella 5.5 placement (**Supplemental Figure 4**).

**Figure 2:**
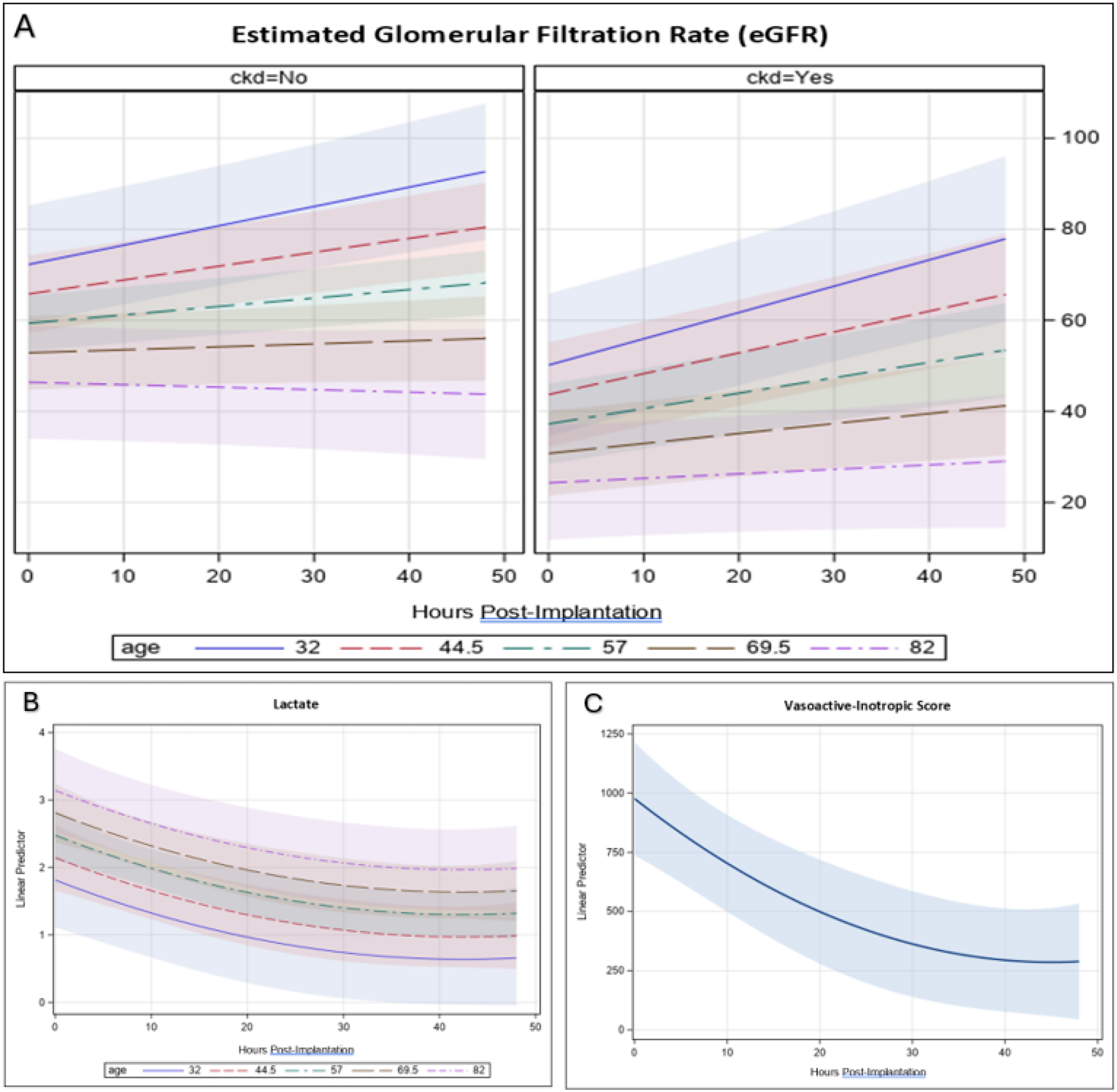
Mixed effects longitudinal models of changes in hepatorenal function and end-organ perfusion after Impella 5.5 implantation. (A) Estimated glomerular filtration rate (eGFR) improved in all patients, although those with pre-existing chronic kidney disease (CKD) experienced the greatest benefit. (B-C) Lactate and vasopressor-inotropic score both decreased. Older age was found to correlate with lower eGFR and higher lactate at time of Impella implantation and a more attenuated response. Bands indicate 95% confidence intervals.

### Clinical course and outcomes

In our cohort, 22 patients died during index hospitalization, 17 received an LVAD, and 33 underwent HT. In total, 8 patients developed severe RV failure after LVAD and only 1 patient developed RV-PGD after HT. The clinical course of our CS cohort, stratified by baseline SCAI staging, is shown in **Figure 3** and **Supplemental Table I**, with 57% of patients bridged to heart replacement therapies.

**Figure 3:**
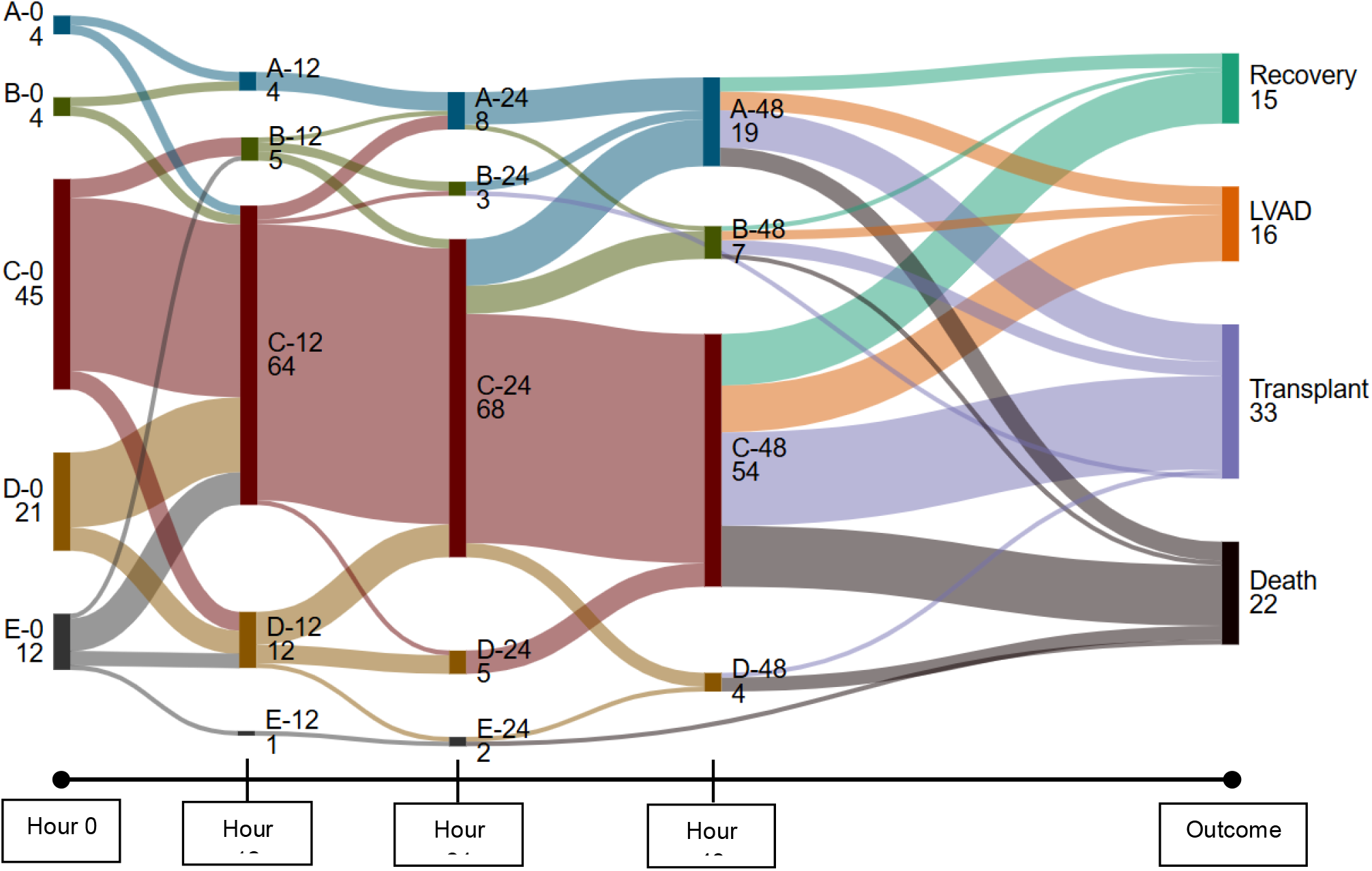
Sankey diagram of Society for Cardiovascular Angiography & Interventions (SCAI) staging of cardiogenic shock patients after Impella 5.5 implantation over the first 48 hours and their clinical outcome. Over a quarter of patients died during index hospitalization and 57% of patients were bridged to left ventricular assist device (LVAD) or heart transplant. Made with SankeyMATIC.

In multivariable logistic regression analysis, age and baseline RV function did not predict the primary and secondary outcomes. Between pre-implantation and 48 hours post-Impella 5.5, each standard deviation increase in RA/PCWP was associated with death (OR 2.34, CI 1.10-6.30, p=0.025) and the composite of death, post-LVAD severe RV failure, and post-HT RV-PGD (OR 2.78, CI 1.35-7.47, p=0.003, **Figure 4**). Additional pre-specified sensitivity analyses were performed, incorporating sex and the slope of change between pre-implantation, 12, 24, and 48 hours after Impella 5.5 to better account for the overall rate of change. In our sensitivity analyses, RA/PCWP remained prognostic and was associated with 2.36-fold higher rate of death (CI 1.09-6.35, p=0.028) and 2.50-fold higher rate of death, post-LVAD severe RV failure, and post-HT RV-PGD (CI 1.27-6.00, p=0.007, **Supplemental Figure 5**). Sex and rates of change in RAP, PAPi, RVSWI, and Ea were not predictive of our clinical outcomes.

**Figure 4:**
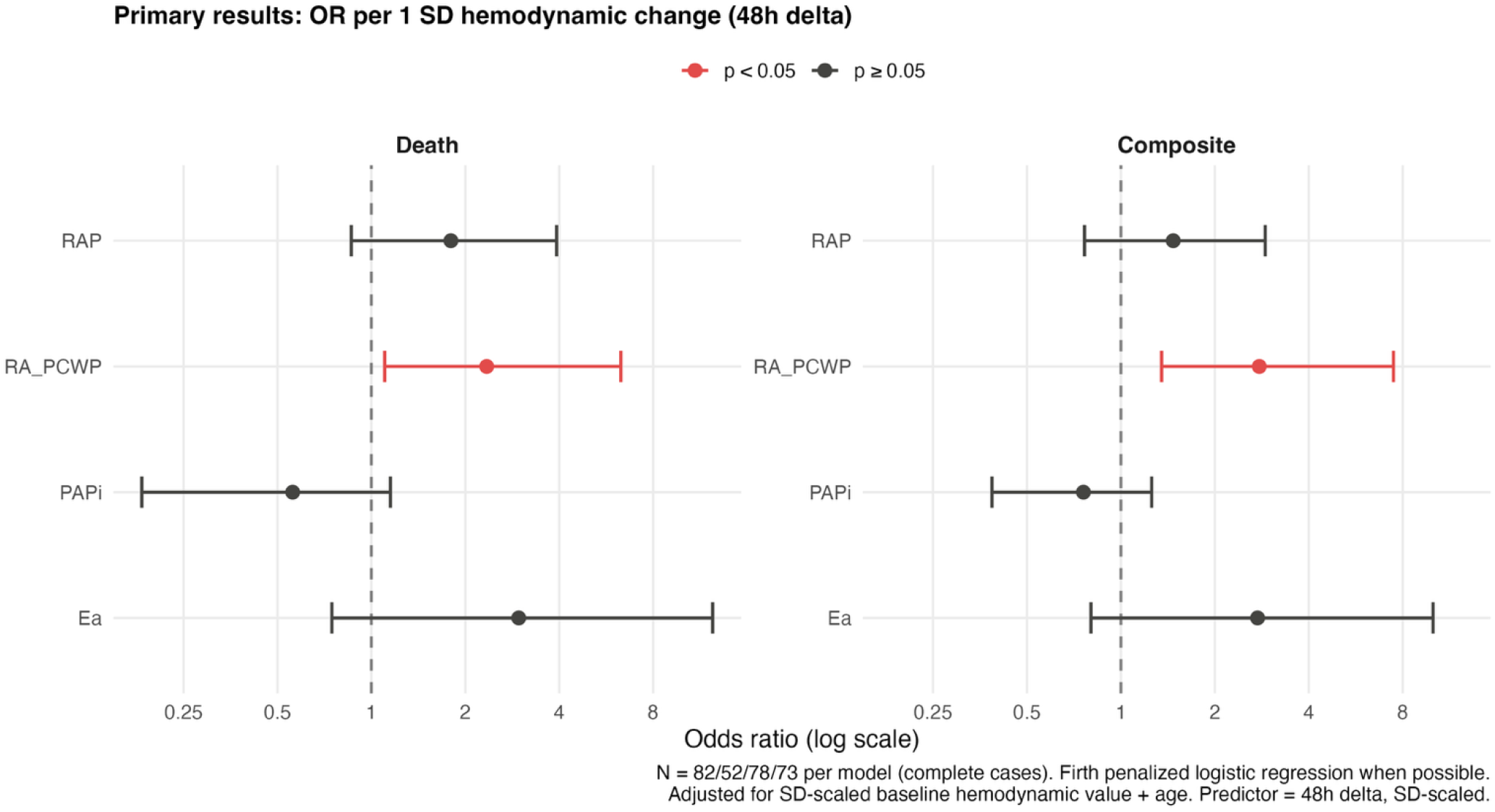
Cox proportional hazards model for death and composite of death, severe right ventricular (RV) failure after left ventricular assist device, and post-transplant RV-primary graft dysfunction. Our primary model adjusted for age, baseline RV function, and the rate of change in hemodynamic variables. Only an increase in right atrial:pulmonary capillary wedge pressure (RA/PCWP) ratio was independently associated with the adverse outcomes. RAP=right atrial pressure,, PAPi=pulmonary artery pulsatility index, Ea=pulmonary artery elastance. Odds ratios and 95% confidence intervals shown.

## Discussion

We found that patients with AMI-CS and HF-CS experienced net improvements in hemodynamics and end-organ perfusion after Impella 5.5 placement, even in those with marginal baseline RV function. RV dysfunction itself did not predict clinical outcomes; however, an increase in RA/PCWP despite tMCS support predicted death and composite of death, post-LVAD RV failure, and post-HT RV-PGD. Our study sheds important insight into expected responses after Impella 5.5 to help inform patient selection for tMCS and identifies the response of RA/PCWP as a key prognosticator in CS after Impella 5.5 implantation.

Despite increasing utilization of Impella 5.5 to treat CS and to bridge patients to heart replacement therapies, few studies have described the anticipated hemodynamic alterations, especially in those with pre-existing RV dysfunction. RV dysfunction is highly prevalent in CS and is an independent predictor of adverse outcomes.^7,8^ As such, when choosing to implant a univentricular tMCS device, clinicians must carefully balance the benefit of tMCS against the harm of added preload to an already vulnerable RV.^16,17^ In this study, we found that Impella 5.5 placement led to a reduction in RAP and improvement in PAPi and RVSWI across all RV function subtypes. In fact, patients with the worst baseline RV function experienced the greatest fall in RAP. We suspect that by offloading the pulmonary circulation and improving renal perfusion, Impella 5.5 rapidly improved filling pressures, especially for those with the greatest congestion. On the other hand, patients with bad RV function exhibited a more modest improvement in PAPi and RVSWI. These are markers of intrinsic RV function and thus the ability for Impella 5.5 to augment PAPi and RVSWI are more attenuated. Importantly, the peak effect was seen within 36 hours after implantation, suggesting that the bulk of hemodynamic benefit occurs early. Overall, our results suggest that the improved forward flow and afterload-reducing effects (lower Ea) of Impella 5.5 is a net-benefit and that CS patients should not be excluded from Impella 5.5 based on RV dysfunction alone.

Interestingly, we identified sex as an independent predictor of RA/PCWP and Ea with women demonstrating more dynamic increase in RA/PCWP and decline in Ea. These results may be related to physiologic sex-differences as women with pulmonary arterial hypertension were found to have smaller RV volumes offset by greater contractility to maintain appropriate RV-PA coupling.^18,19^ Similar to a study by Asher et al, we did not find sex to be an independent predictor of mortality, although further investigation into sex-specific differences in RV-PA coupling after tMCS is necessary.^20^

In addition to improving the hemodynamic profile, Impella 5.5 also led to better end-organ perfusion, with rapid decrease in lactate and VIS and increase in eGFR. First, we noted that all patients, particularly those with pre-existing CKD, experienced significant improvement in renal function after Impella 5.5 implantation. We believe these patients had under-appreciated cardiorenal syndrome, which occurs in ~30% of CS cases, and the rapid fall in RAP and restoration of forward flow partially reversed the pathophysiology.^21^ Second, we found an age-dependent effect, with older patients having higher lactate and lower eGFR at baseline and experiencing a significantly more attenuated benefit after Impella 5.5 placement (**Figure 2a-b**). This could be due to age-related changes in cardiac and renal reserve from expected decline in diastolic function, nephron number, and renal blood flow.^22,23^ While our findings do not support withholding Impella 5.5 due to age alone, future studies should investigate the mechanisms behind this finding. Lastly, we did not find any changes in hepatic function as ascertained by MELD, likely due to the short timeframe of this study as bilirubin changes often lag behind other liver-specific biomarkers.

Importantly, we found that baseline RV dysfunction in AMI and HF-CS patients supported by Impella 5.5 was not associated with clinical outcomes. However, a detrimental increase in RA/PCWP despite tMCS support was predictive of death and the composite of death, post-LVAD RV failure, and post-HT RV PGD, respectively. The syndrome of right heart failure is a systemic process and elevated filling pressures lead to splanchnic congestion, release of inflammatory cytokines, and end-organ dysfunction.^10,24,25^ RAP has consistently been shown to predict mortality across ambulatory, hospitalized, and advanced HF disease states, although it was not associated with outcomes in our cohort.^15,26^ Instead, we found that while RAP improved in all RV dysfunction subgroups after Impella 5.5, RA/PCWP paradoxically increased. This was driven by a more significant reduction in PCWP due to robust LV unloading (preferentially increasing the RA/PCWP ratio). Thus, we hypothesize that RA/PCWP is an even more sensitive and prognostic marker than RAP and can differentiate patients who respond favorably to univentricular tMCS.^15^ This is consistent with multicentered studies showing RA/PCWP is a predictor of severe RV failure and mortality after durable LVAD.^27–29^ Furthermore, our study supports serial hemodynamic measurements to assess response to treatment in CS and initial values, no matter how grim, should not by itself preclude prompt tMCS support.^30^ However, in CS patients who exhibit unfavorable response in RA/PCWP despite Impella 5.5 support, clinicians should consider adding mechanical RV support, initiating earlier evaluation for heart replacement therapies, or involving palliative care given the negative prognosis.

### Limitations

This study has several limitations. First, this is a single-centered study and our results may not be generalizable to other institutions. However, our study carefully describes anticipated hemodynamic changes after Impella 5.5 and we believe this will be of interest to the HF and CS community. Second, we studied acute hemodynamic changes within 48 hours but acknowledge that further alterations are possible beyond this initial timeframe. We chose the early timepoints because of the critical “golden hours” of CS, during which early support is critical for therapeutic benefit. Future research should focus on longer follow up to assess how durable these initial hemodynamic changes are. Third, we excluded patients who underwent concurrent ECMO or RV tMCS, resulting in selection bias as patients with the greatest RV dysfunction may not have been implanted with Impella 5.5 in the first place. Fourth, we focused on specific hemodynamic parameters but acknowledge that other variables may contribute to mortality in CS and RV dysfunction is not fully captured by TAPSE and RA/PCWP alone. Finally, given that Impella 5.5 was used as a bridge to LVAD or HT in >50% of our cohort, the clinical outcomes may have been influenced by these subsequent treatments or unmeasured factors (e.g. social support, etc), which this study is not designed to account for. Nevertheless, tMCS decisions in critically ill CS patients are frequently made without full understanding of patients’ candidacy for heart replacement therapies and our study reflects current clinical practice.

## Conclusion

In conclusion, we found that Impella 5.5 improved RV-PA hemodynamics and systemic perfusion in patients with AMI and ADHF-CS. RV dysfunction itself should not preclude the use of univentricular left-sided support with Impella 5.5 and an increase in RA/PCWP despite tMCS is a powerful prognosticator for death and poor clinical outcomes.

## Data Availability

All data produced in the present study are available upon reasonable request to the authors.

