## Supplemental material for "Acute hemodynamic effects after Impella 5.5 in cardiogenic shock and association with clinical outcomes"

**Supplemental Materials**

**Detailed Analyses of Change in Hemodynamics and Markers of Hypoperfusion**

Due to our findings of curvilinear (quadratic) effects over time for many of the hemodynamic parameters and markers of hypoperfusion we analyzed, we were sometimes limited to verbally describing trends over time or displaying graphical illustrations of them rather than reporting rate changes which varied across time. Most dynamic change was seen immediately post-implantation with effects typically appearing to plateau at around 30 hours post implantation and sometimes even reversing direction slightly as time wore on (although the latter may be partly an artifact of the quadratic function fit and the longer time interval between the third and final assessments).

**Hemodynamic Parameters**

**Right atrial pressure**

We found a significant interaction of RV Function with quadratic time (p = 0.0004), demonstrating a decline in RAP across all RV subgroups but most dramatically seen in those with poor RV function. (See Figure 1(A)). About 22% of the variance in RAP was accounted for by model fixed effects. There were no significant effects involving age or sex. Model residuals from RAP values predicted by fixed and random effects reasonably conformed to normality assumptions, as was the case for all analyses reported below.

**RA:PCWP**

Conversely, in the analysis of RA:PCWP, we see a paradoxical increase across all RV subgroups. This was due to a more significant reduction in PCWP due to Impella 5.5’s LV unloading effects. There was also a significant interaction of sex and quadratic time, wherein females showed a somewhat bell-shaped mean trajectory across time whereas the males mean trajectory demonstrated a more gradual and linear incline (p = 0.0009; see Supplemental Figure 1). Orthogonal to this, there was also a significant main effect of RV Function (p = 0.0448). Post hoc analysis with Tukey adjustments of p values showed that this effect reflected the fact that the Bad and Intermediate groups each had a marginally significantly (p≤0.06) higher mean RA:PCWP across time than the Good group. There were no significant effects involving age.

**Pulmonary artery pulsatility index**

We found a significant quadratic time effect (p = 0.026) and an interaction of RV Function with the linear component of time (p = 0.015), reflecting the fact that the Bad and Intermediate groups showed a somewhat gradual increase in mean PAPI across time leveling at about 30 hours post implantation whereas the Good group started higher and showed an essentially linear further increase throughout the time period (see Figure 1(B)). There were no significant effects involving age or sex. About 10% of variance in PAPI was accounted for by the model fixed effects.

**Pulmonary artery elastance (E_a_)**

We found a significant quadratic effect of time in which E_a_ values declined sharply initially and then leveled off around 30 hours (p < 0.001). There was also a significant interaction of sex with quadratic time (p = 0.0035), wherein females showed a more dramatic decline than males. RV function also had a significant interaction with linear time (p = 0.0086), in which the Good group also showed stronger evidence of the above relation across time than the other groups. (See Supplemental Figure 2). There was no significant effect involving age. Model fixed effects accounted for a third of the variance in Ea.

**RV Stroke Work Index**

We found a significant interaction of RV function with quadratic time (p = 0.0188), in which the Bad and Intermediate groups showed a gradual linear increase in mean trajectory across time whereas the Good group showed a more rapid initial increase before leveling off at about 30 hours post implantation (see Figure 1(C)). There were no significant effects involving age or sex. Model fixed effects accounted for 9% of the variance in RVSWI.

**Markers of Hypoperfusion**

**eGFR**

We found a significant interaction of CKD and linear time, demonstrating a steeper rise by 0.15 units/hr (95% CI: 0, 0.31; p=0.05) in eGFR in those with history of CKD as compared to those without. On average, patients with CKD had an increase of 0.88 units/hr (95% CI: 0.48,1.28; p < 0.0001). There was also a significant interaction of age with linear time, revealing that older patients experienced a slight attenuation in their increase over time in eGFR compared to younger counterparts (p = 0.003). (See Figure 2(A)). About 23% of the variance in eGFR was accounted for by model fixed effects.

**Lactate**

The quadratic time main effect was significant (p = 0.05), reflecting a decline across time before plateauing at around 30 hours post implantation. Orthogonal to this effect, there was also a significant main effect of age with older people having higher lactate values across time as a whole than younger (by 0.027 Lactate units per year of age (95% CI: 0.004 0.049; p=0.02) (See Figure 2(B)). Fixed effects accounted for about 8% of the variance of Lactate.

**Vasoactive-Inotropic Score**

The quadratic time main effect was significant (p = 0.0466), reflecting a decline across time leveling off around 40 hours post implantation. No other fixed effects were significant. (See Figure 2(C)). Fixed effects accounted for about 5% of the variance of the Vasoactive-Inotropic Score.

**MELD score**

Unlike the longitudinal analyses above, the time predictor for MELD was binary (Pretest vs. 48-hour post-test). There was no significant change in MELD across time. However, there were significant main effects for age (p=0.016) and for CKD (p=0.0023). Age had a positive relation to MELD with an average rise in MELD of 0.13 (SE=0.05) units per year of age. As time was nonsignificant, Figure 3 shows predicted MELD scores across age in years rather than time as the effect of the latter was predicted to be flat. Patients with CKD had a significantly higher adjusted MELD mean (p = 0.0023) by 3.87 (SE=1.23) units, which was not unexpected as creatinine is a component of MELD scoring. About 11% of the variance in MELD was predicted by model fixed effects.

***Supplemental Tables and Figures***

**Supplemental Table I: SCAI Stage Transitions After Impella 5.5**. SCAI shock stage distribution at each time point after Impella 5.5 implantation, stratified by clinical outcome. Values are n (% of outcome group). *Composite = death, post-LVAD severe RV failure, or post-transplant RV-PGD, excluding patients counted under Death. SCAI = Society for Cardiovascular Angiography & Interventions; LVAD = left ventricular assist device; PGD = primary graft dysfunction.

| **Time Point** | **SCAI Stage** | **Death n = 22** | **Composite* n = 9** | **Survived / No event** |
| --- | --- | --- | --- | --- |
| **Pre-implant** | **A** | **0 (0.0%)** | **1 (11.1%)** | **3 (5.5%)** |
|  | **B** | **2 (9.1%)** | **0 (0.0%)** | **2 (3.6%)** |
|  | **C** | **5 (22.7%)** | **5 (55.6%)** | **35 (63.6%)** |
|  | **D** | **7 (31.8%)** | **3 (33.3%)** | **11 (20.0%)** |
|  | **E** | **8 (36.4%)** | **0 (0.0%)** | **4 (7.3%)** |
|  | **Total** | **22** | **9** | **55** |
| **12 hours** | **A** | **1 (4.5%)** | **0 (0.0%)** | **3 (5.5%)** |
|  | **B** | **1 (4.5%)** | **1 (11.1%)** | **3 (5.5%)** |
|  | **C** | **13 (59.1%)** | **7 (77.8%)** | **44 (80.0%)** |
|  | **D** | **6 (27.3%)** | **1 (11.1%)** | **5 (9.1%)** |
|  | **E** | **1 (4.5%)** | **0 (0.0%)** | **0 (0.0%)** |
|  | **Total** | **22** | **9** | **55** |
| **24 hours** | **A** | **3 (13.6%)** | **0 (0.0%)** | **5 (9.1%)** |
|  | **B** | **1 (4.5%)** | **1 (11.1%)** | **1 (1.8%)** |
|  | **C** | **14 (63.6%)** | **7 (77.8%)** | **47 (85.5%)** |
|  | **D** | **2 (9.1%)** | **1 (11.1%)** | **2 (3.6%)** |
|  | **E** | **2 (9.1%)** | **0 (0.0%)** | **0 (0.0%)** |
|  | **Total** | **22** | **9** | **55** |
| **48 hours** | **A** | **4 (19.0%)** | **2 (22.2%)** | **13 (24.1%)** |
|  | **B** | **1 (4.8%)** | **0 (0.0%)** | **6 (11.1%)** |
|  | **C** | **13 (61.9%)** | **7 (77.8%)** | **34 (63.0%)** |
|  | **D** | **3 (14.3%)** | **0 (0.0%)** | **1 (1.9%)** |
|  | **E** | **0 (0.0%)** | **0 (0.0%)** | **0 (0.0%)** |
|  | **Total** | **21** | **9** | **54** |

**Supplemental Figure 1: Consort diagram of cardiogenic shock (CS) patients supported with Impella 5.5 between 2019-2023.** We included patients with acute myocardial infarction (AMI) and acute decompensated heart failure (HF)-induced CS. We excluded those who underwent high-risk procedures such as ventricular tachycardia ablation, percutaneous coronary intervention, and coronary artery bypass graft surgery and those with concurrent extracorporeal membrane oxygenation (ECMO) or right ventricular assist device (RVAD).
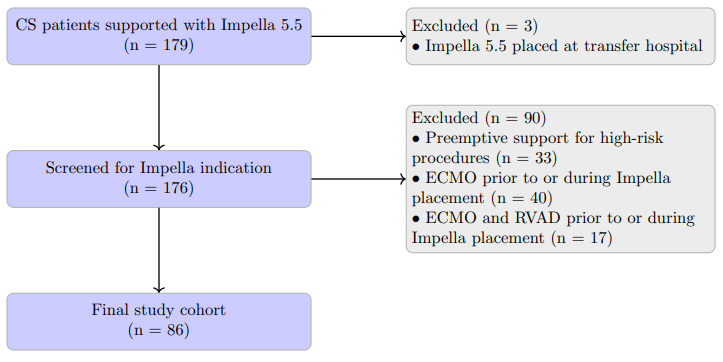

**Supplemental Figure 2: Mixed effects longitudinal model for right atrial:pulmonary capillary wedge pressure (RA/PCWP) ratio following Impella 5.5 placement, stratified by good, intermediate, and bad right ventricular (RV) function.** Bands indicate 95% confidence intervals.

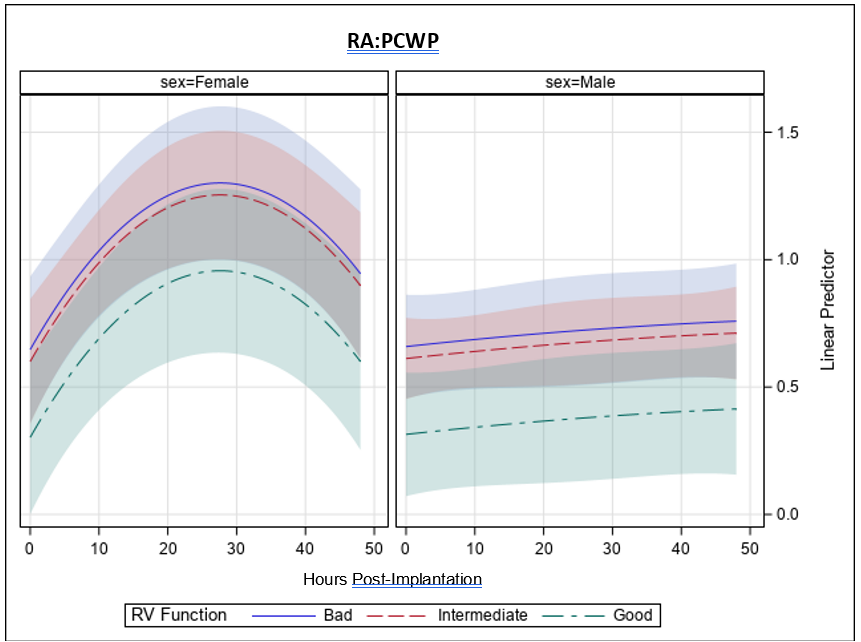

**Supplemental Figure 3: Mixed effects longitudinal model for pulmonary artery elastance following Impella 5.5 placement, stratified by good, intermediate, or bad right ventricular (RV) function.** Bands indicate 95% confidence intervals.

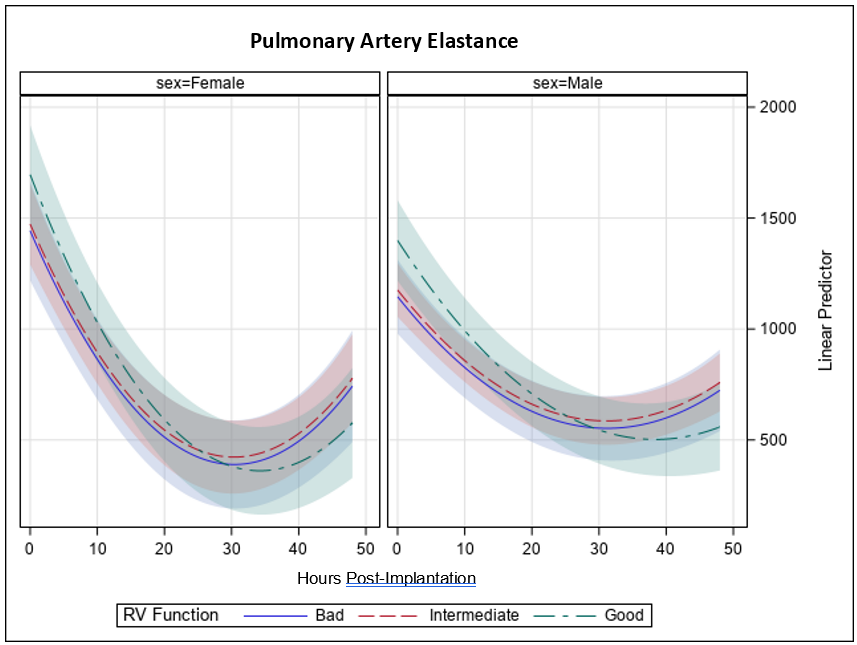

**Supplemental Figure 4: Mixed effects longitudinal model for Model for end-stage liver disease (MELD) after Impella 5.5 placement, stratified by history of CKD.** Bands indicate 95% confidence intervals.

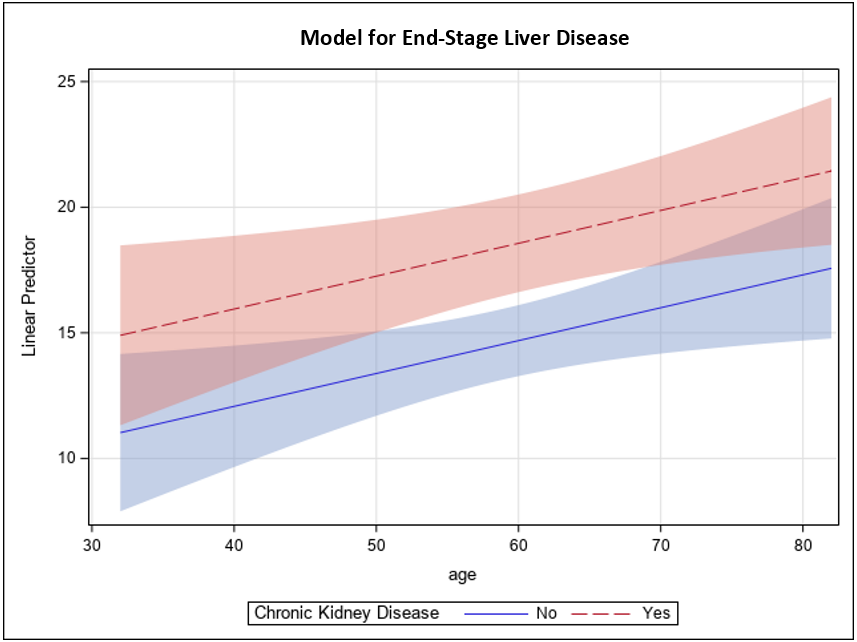

**Supplemental Figure 5: Cox proportional hazards model for death and composite of death, severe right ventricular (RV) failure after left ventricular assist device, and post-transplant RV-primary graft dysfunction.** Pre-specified sensitivity analysis adjusted for age, baseline RV function, and linear regression of the slope of change between pre-implantation, 12, 24, and 48 hours after Impella 5.5. Right atrial:pulmonary capillary wedge pressure (RA/PCWP ratio) is associated with both clinical outcomes. RAP=right atrial pressure, RA/PCPW=right atrial-pulmonary capillary wedge ratio, PAPi=pulmonary artery pulsatility index, Ea=pulmonary artery elastance. Odds ratios and 95% confidence intervals shown.
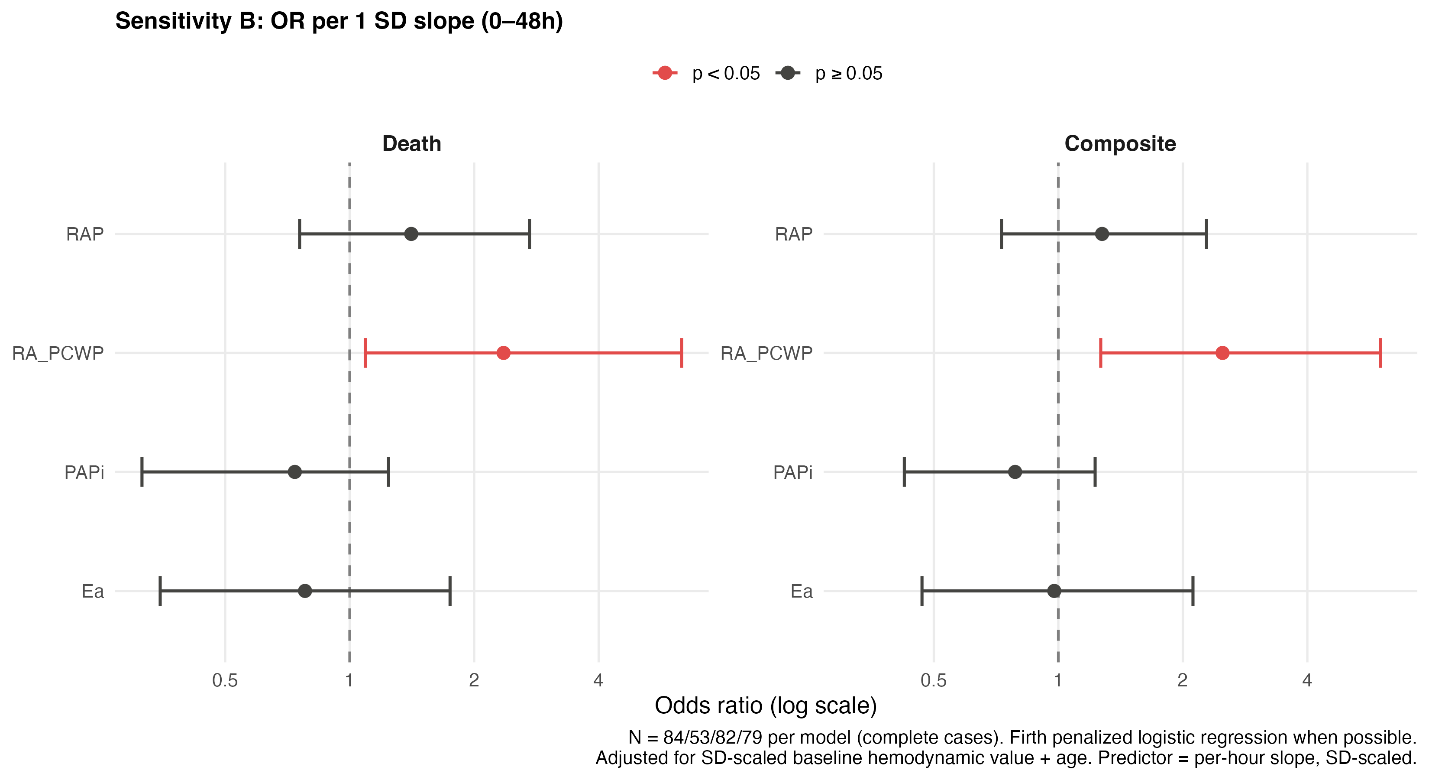
